# Total nodule number is an independent prognostic factor in resected stage III non-small cell lung cancer: a deep learning powered study

**DOI:** 10.1101/2021.03.03.21252811

**Authors:** Xiuyuan Chen, Qingyi Qi, Zewen Sun, Dawei Wang, Jinlong Sun, Weixiong Tan, Xianping Liu, Taorui Liu, Nan Hong, Fan Yang

**Affiliations:** Department of Thoracic Surgery, Peking University People’s Hospital, Beijing, China; Department of Radiology, Peking University People’s Hospital, Beijing, China; Institute of Advanced Research, Beijing Infervision Technology Co., Ltd., Beijing, China

**Keywords:** nodule number, non-small cell lung cancer, prognosis, artificial intelligence, multiple pulmonary nodules

## Abstract

Almost every lung cancer patient has multiple pulmonary nodules while the significance of nodule multiplicity in locally advanced non-small cell lung cancer (NSCLC) remained unclear. This study explores the relationship between deep learning detected total nodule number (TNN) and survival outcomes in patients with surgical resected stage I-III NSCLC. Patients who underwent surgical resection for stage I-III NSCLC with accessible preoperative chest CT scan from 2005 to 2018 were identified from our database. Deep learning-based AI algorithms using convolutional neural networks (CNN) was applied for pulmonary nodule (PN) detection and classification. Of the 2126 patients, a total number of 33410 PNs were detected by AI. Median TNN detected per person was 12 (IQR 7-20). AI-detected TNN (analyzed as continuous variable) was independent prognostic factor for both RFS (HR 1.012, 95% CI 1.002-1.022, p = 0.021) and OS (HR 1.013, 95% CI 1.002-1.025, p = 0.021) in multivariate analyses of stage III cohort; while it was not significantly associated with survival in stage I and II cohorts. In terms of nodule categories, the numbers of upper-lobe nodule, same-side nodule, other-side nodule, solid nodule, and even solid nodule at small size (≤ 6mm) were independent prognostic factors; while the numbers of middle/lower-lobe nodule, same-lobe nodule, subsolid nodule, calcific nodule and perifissural nodule were not associated with survival. In survival tree analysis, rather than using traditional IIIA and IIIB classification, the model grouped cases by AI-detected TNN (lower vs. higher: log-rank p < 0.001), which showed superior discrimination of survival in stage III cohort. In conclusion, AI-detected TNN was significantly associated with survival in patients with surgical resected stage III NSCLC. Lower TNN detected on preoperative CT scan indicated better prognosis in patients who underwent complete surgical resection.

## INTRODUCTION

Lung cancer is a leading cause of cancer-related death worldwide [1]. Since early detection of cancer is an important opportunity for decreasing mortality, multiple randomized trials and guidelines recommend lung cancer screening using low-dose CT (LDCT) for high-risk individuals [2-7]. With the adoption of LDCT for lung cancer screening, the number of chest CT scans increase dramatically each year [8]. To address the repetitive and burdensome work of dealing with mostly normal images, computer aided detection/diagnosis (CAD), which has a computer to perform a given task consistently and tirelessly, becomes extremely appealing [9].

CAD supported by machine-learning techniques has been utilized to detect PN since 2002 [10]. Although standardized CAD systems have been proven to improve diagnostic accuracy, few of them have been implemented in actual clinical practice due to its high dependence on imaging processing and false positive rates [11, 12]. In recent years, deep-learning based AI algorithms using convolutional neural networks (CNN) have attracted considerable attention in the area of machine-learning. The key advantage that CNNs have over conventional CAD techniques is their ability to self-learn previously unknown features, maximizing classification with limited direct supervision [13]. Thus, CNNs bring about a significant false-positive reduction in PN detection, recognition, segmentation, and classification [14-19].

The key issue in the management of incidental PNs detected on CT images is to differentiate benign and malignant nodules. Radiological features, such as larger nodule size, upper lobe location, marginal spiculation, and faster growth rate are generally considered as risk factors for malignancy [20-25]. These principles mainly focus on the assessment of the largest or most suspicious nodule. However, half of the patients detected with PNs have multiple nodules [26]. Nodule multiplicity, which is a potential indicator for malignancy, is commonly overlooked. Only limited data concerning the relationship between TNN and lung cancer probability is available. In the PanCan and BCCA trials, lower TNN was associated with an increased risk of lung cancer [20]. While another study analyzing patients from the NELSON trial showed that the risk of lung cancer increased as TNN rising from 1 to 4, but decreased in patients with 5 or more nodules [26].

The results of above-mentioned screening trials indicated that TNN was either negatively or not significantly associated with lung cancer probability, which might reflect a low incidence of multiple malignancies in screening population [27]. However, for patients with high pretest probability of malignancy, whether TNN plays a role in determining lung cancers probability with multiple pulmonary sites of involvement, distinguishing multiple primary lung cancers (MPLC) from intrapulmonary metastasis (IPM), and thus, predicting prognosis remains unknown. The purpose of this study was to calculate TNN detected on preoperative CT images using CNN-based AI algorithm, and explore in-depth the relationship between AI-detected TNN and survival outcomes in patients with resectable stage I-III NSCLC.

## MATERIALS AND METHODS

### Patients

We retrospectively reviewed the medical records of patients pathologically diagnosed with stage I-III (according to the 8^th^ edition of American Joint Committee on Cancer [AJCC] prognostic group) NSCLC who underwent surgical resection at the department of thoracic surgery of Peking University People’s Hospital from October 2005 to December 2018. Only patients received preoperative chest CT scan within 90 days prior to the surgery in our institution were included. Patients were excluded: 1) if they received neoadjuvant therapy; 2) if the surgical margin was positive; 3) if perioperative death occurred within 30 days; 4) if the follow-up information was inadequate.

### AI-Powered PN Detection

InferRead™ CT Lung, a wildly used deep learning-based AI algorithm developed by InferVision, was applied for PN detection in this study. Only the last chest CT scan before surgery was chosen. Firstly, PNs was detected by AI algorithm and TNN was calculated accordingly. Then, PNs were classified according to their lobar distributions (left lower lobe, left upper lobe, right lower lobe, right middle lobe, and right upper lobe), locations (same lobe as the primary tumor [same-lobe], ipsilateral lobe different from the primary tumor [same-side], and contralateral lobe [other-side]), and types (solid nodule, mixed ground-glass nodule [m-GGN], pure ground-glass nodule [p-GGN], calcific nodule, and perifissural nodule). Moreover, solid and subsolid (m-GGN and p-GGN) nodules were further categorized based on their size.

### Statistical Analysis

Continuous variables were presented as median with interquartile range (IQR) and were analyzed by using Wilcoxon rank-sum test and one-way analysis of variance (ANOVA). Categorical variables were presented as frequencies and percentages. Survival curves were compared using Kaplan-Meier method with log-rank test. Univariate Cox proportional hazards models were built first to determine which factors were significantly associated with survival. Then, multivariate Cox models were constructed incorporating factors with p ≤ 0.10 identified in the univariate analyses.

In stage III cohort, maximally selected log-rank statistics were used to determine the optimal cutoff value of nodule number for predicting OS. Patients were then dichotomized into lower- and higher-nodule number groups according to the estimated cut-point. Furthermore, least absolute shrinkage and selection operator (LASSO) Cox regression model with cross-validation was utilized to select the most useful prognostic features among all categories of AI-detected nodule numbers.

Finally, survival tree analysis was conducted to generate a tree-based model for survival data using log-rank test statistics for recursive partitioning. This tree-based model grouped cases according to the best split of OS using AI-detected TNN and the 8^th^ edition of AJCC prognostic group. A candidate grouping scheme was then developed based on this tree analysis.

All the statistical analyses were executed using R version 4.0.0 for Windows (R Foundation for Statistical Computing, Vienna, Austria). All the statistical tests were two-sided and p values of0.05 or less were considered statistically significant.

### Ethics

The study involving human participants were reviewed and approved by the Institutional Review Board of Peking University People’s Hospital (2020PHB385-01). Since only de-identified data were used in this study, informed consents for participants were waived by the committee.

## RESULTS

### Characteristics of Patients and Nodules

A total of 2126 patients who underwent surgical resection for stage I-III NSCLC with accessible preoperative chest CT scan were included in this study. The median follow-up time was 33 months (IQR 21-48). The demographic and clinicopathologic characteristics of the patients are summarized in Table 1.

**Table 1.** Characteristics of the Patient Cohort (N = 2126)

| Variables | Value |
| --- | --- |
| Age, y |  |
| Median (IQR) | 61 (54-68) |
| Sex |  |
| Male | 998 (46.9%) |
| Female | 1128 (53.1%) |
| Smoking history |  |
| No | 1456 (68.5%) |
| Yes | 670 (31.5%) |
| Comorbid |  |
| No | 850 (40.0%) |
| Yes | 1276 (60.0%) |
| Surgical approach |  |
| VATS | 1997 (93.9%) |
| VATS converted to open | 61 (2.9%) |
| Open | 68 (3.2%) |
| Surgical procedure |  |
| Sublobar resection | 636 (29.9%) |
| Lobectomy | 1419 (66.8%) |
| Sleeve lobectomy | 39 (1.8%) |
| Pneumonectomy | 32 (1.5%) |
| Histologic type |  |
| Adenocarcinoma | 1780 (83.7%) |
| Squamous Cell Carcinoma | 280 (13.2%) |
| Others | 66 (3.1%) |
| Pathologic T Stage |  |
| T1 | 1383 (65.1%) |
| T2 | 579 (27.2%) |
| T3 | 115 (5.4%) |
| T4 | 49 (2.3%) |
| Pathologic N Stage |  |
| N0 | 1765 (83.0%) |
| N1 | 145 (6.8%) |
| N2 | 216 (10.2%) |
| AJCC Stage (8th edition) |  |
| IA1 | 499 (23.5%) |
| IA2 | 515 (24.2%) |
| IA3 | 265 (12.5%) |
| IB | 347 (16.3%) |
| IIA | 53 (2.5%) |
| IIB | 184 (8.7%) |
| IIIA | 213 (10.0%) |
| IIIB | 50 (2.3%) |
| Complication |  |
| No | 2038 (95.9%) |
| Yes | 88 (4.1%) |
| Adjuvant therapy |  |
| No | 1294 (60.9%) |
| Yes | 445 (20.9%) |
| Unknown | 387 (18.2%) |

IQR, interquartile range; VATS, video-assisted thoracoscopic surgery.

The framework of deep-learning powered PN detection algorithm and an example of 3D reconstruction of AI-detected nodules are shown in Figure 1A and B. Of the 2126 patients, a total number of 33410 PNs were detected. The features of these AI-detected nodules are given in Table 2. The distributions of AI-detected TNN, solid nodule number, and subsolid nodule number per person were all positively skewed, and the medians of these three above-mentioned nodules were 12 (IQR 7-20), 6 (IQR 3-10), and 3 (IQR 1-6) respectively (Figure 2A to C).

**Table 2.**
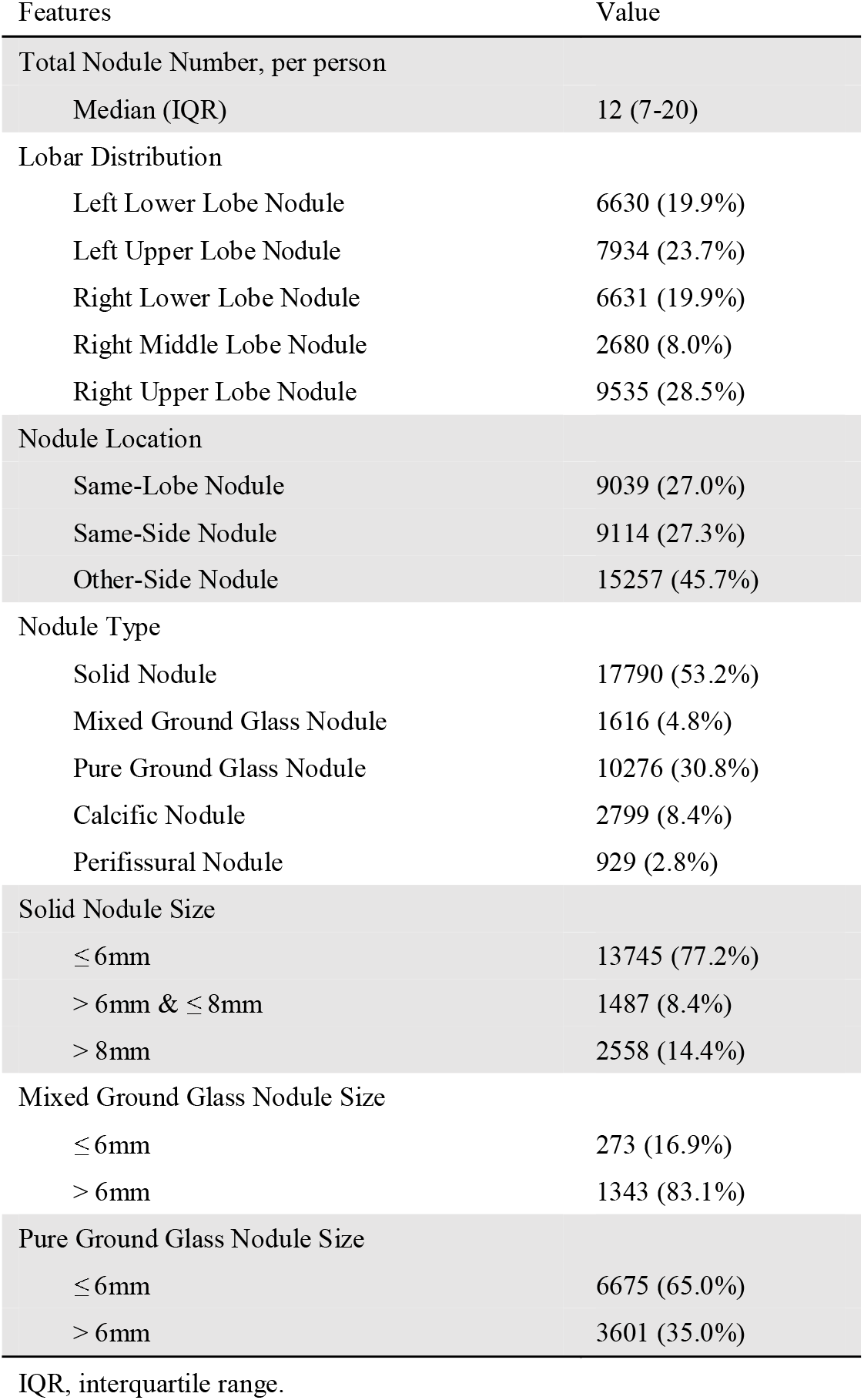
Characteristics of AI-Detected Pulmonary Nodules (n = 33410)

**Figure 1.**
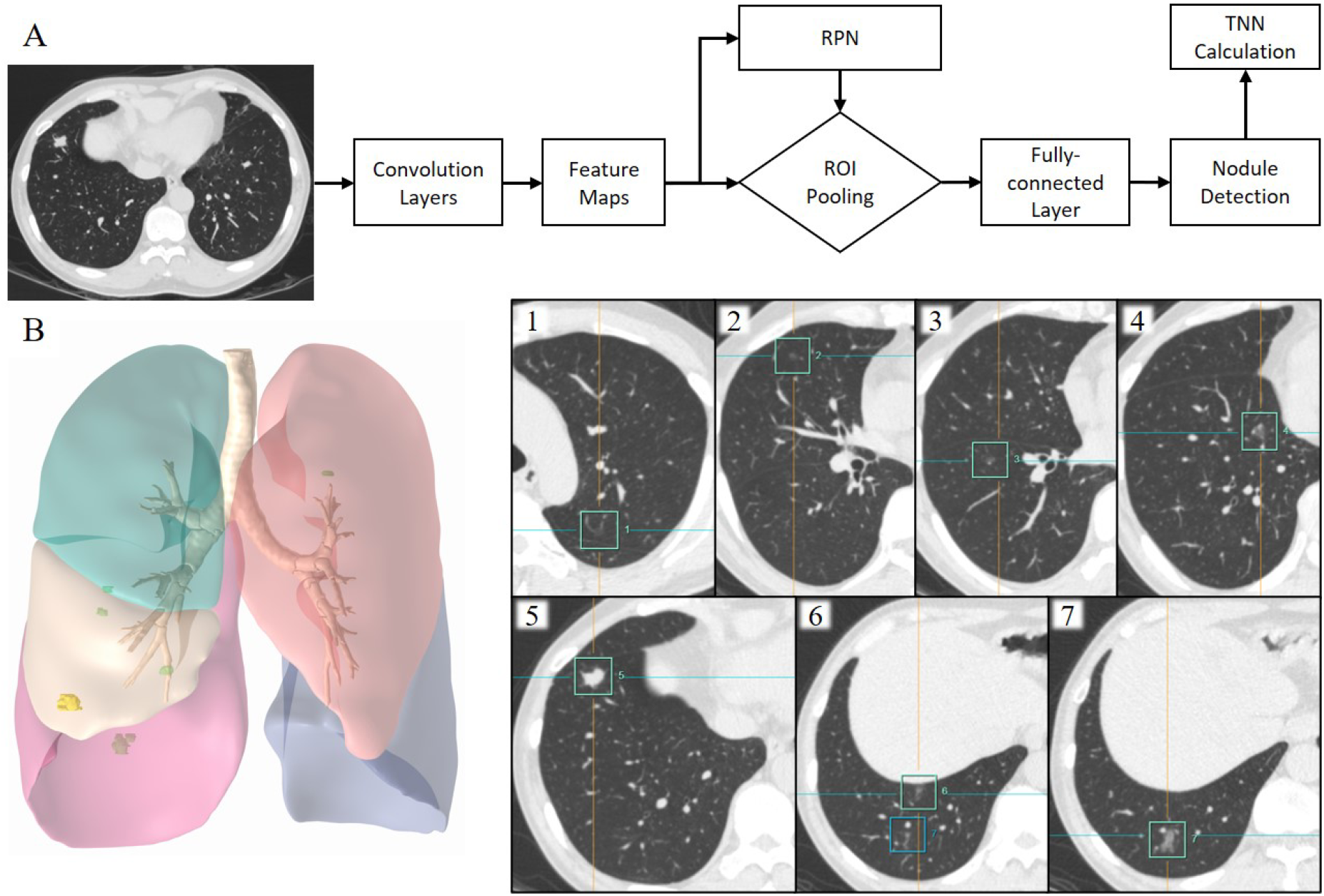
The framework of deep learning powered pulmonary nodule detection algorithm and an example of 3D reconstr uction of AI-detected nodules with corresponding CT images under the lung window setting. (A) Feature maps are extracted using CNN. A RPN is used to obtain potential regions from extracted features. After ROI pooling and fully-connected layers, nodules are detected with rectangular proposals. (B) Seven nodules are detected by AI algorithm, including 1 solid nodule (#5), 2 mixed GGNs (#4, #7), and 4 pure GGNs (#1, #2, #3, #6). RPN, regional proposal network; ROI, region of interest; TNN, total nodule number; CNN, convolutional neural network; GGN, ground-glass nodule.

**Figure 2.**
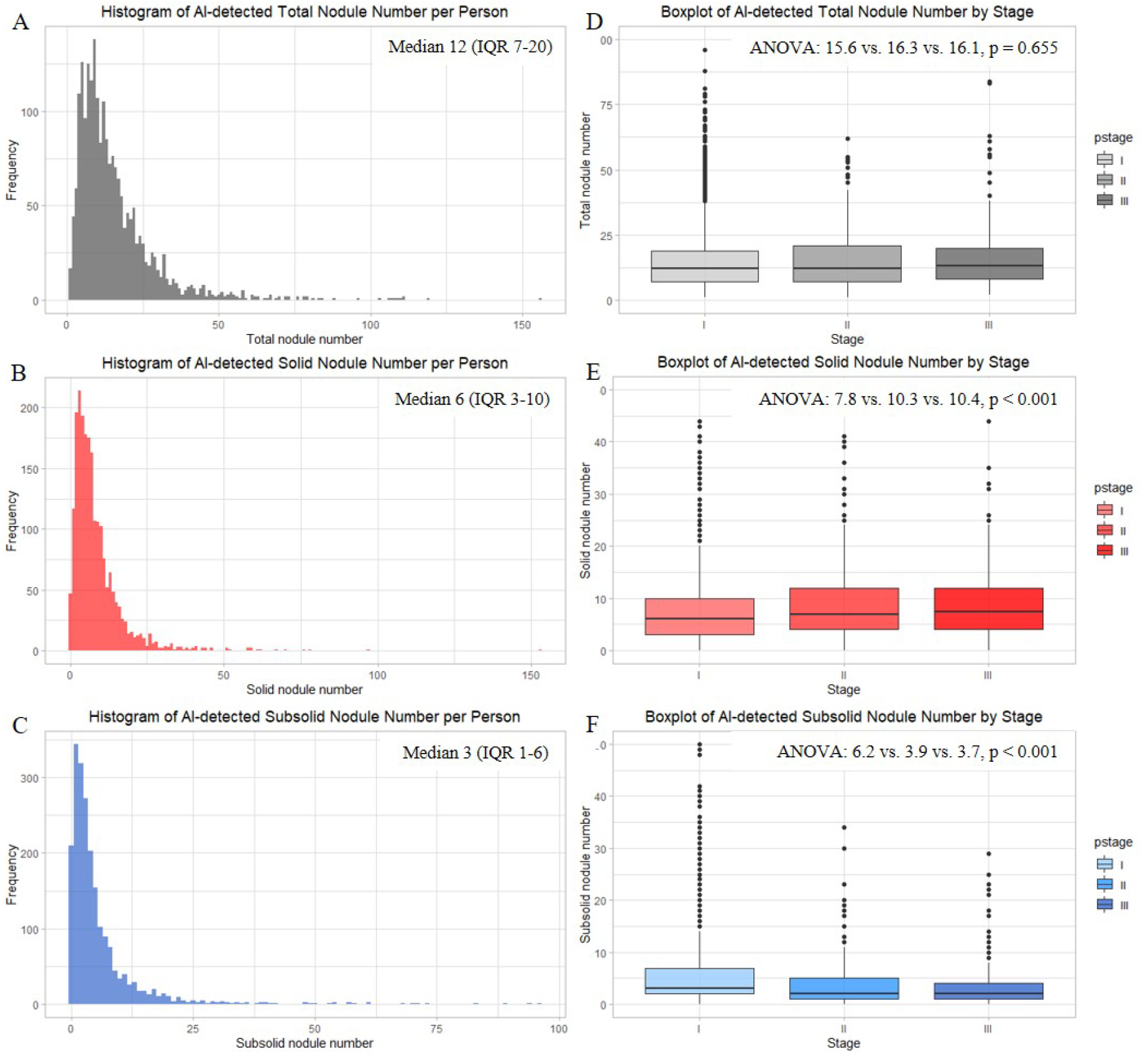
Frequency distribution of AI-detected nodules. (A) TNN, (B) solid nodule number, (C) subsolid nodule number, (D) TNN stratified by pathological stage, (E) solid nodule number stratified by pathological stage, (F) subsolid nodule number stratified by pathological stage. TNN, total nodule number; IQR, interquartile range; ANOVA, analysis of variance.

When considering the discrepancy of nodule number among different stages, we found that there was no statistically significant difference between means of TNN (one-way ANOVA p = 0.655). However, means of solid nodules were significantly higher in patients with stage II and III disease, while means of subsolid ones were higher in those with stage I disease (both p < 0.001, Figure 2D to F). Moreover, patients with late-stage disease tended to have more solid nodules with greater size (Supplementary Figure 1).

### Survival Analyses

We analyzed the survival of the patients by stage according to the 8^th^ edition of AJCC prognostic group (Figure 3A and B). The differences of both RFS and OS between any two stages were statistically significant (pairwise comparison p < 0.001). Cox proportional hazards models were then built to determine the prognostic factors of the entire cohort (Supplementary Table). In univariate analysis, lower AI-detected TNN (as continuous variable) was associated with improved RFS (HR 1.008, 95% CI 1.001-1.014, p = 0.017), while it was not significantly associated with improved OS (HR 1.006, 95% CI 0.999-1.012, p = 0.099). In multivariate analysis, TNN was neither an independent prognostic factor for RFS (HR 1.006, 95% CI 0.999-1.012, p = 0.080) nor for OS (HR 1.002, 95% CI 0.995-1.009, p = 0.590) after adjusting for age, sex, smoking history, surgical approach, surgical procedure, histologic type, adjuvant therapy status, and pathologic T and N stage.

**Figure 3.**
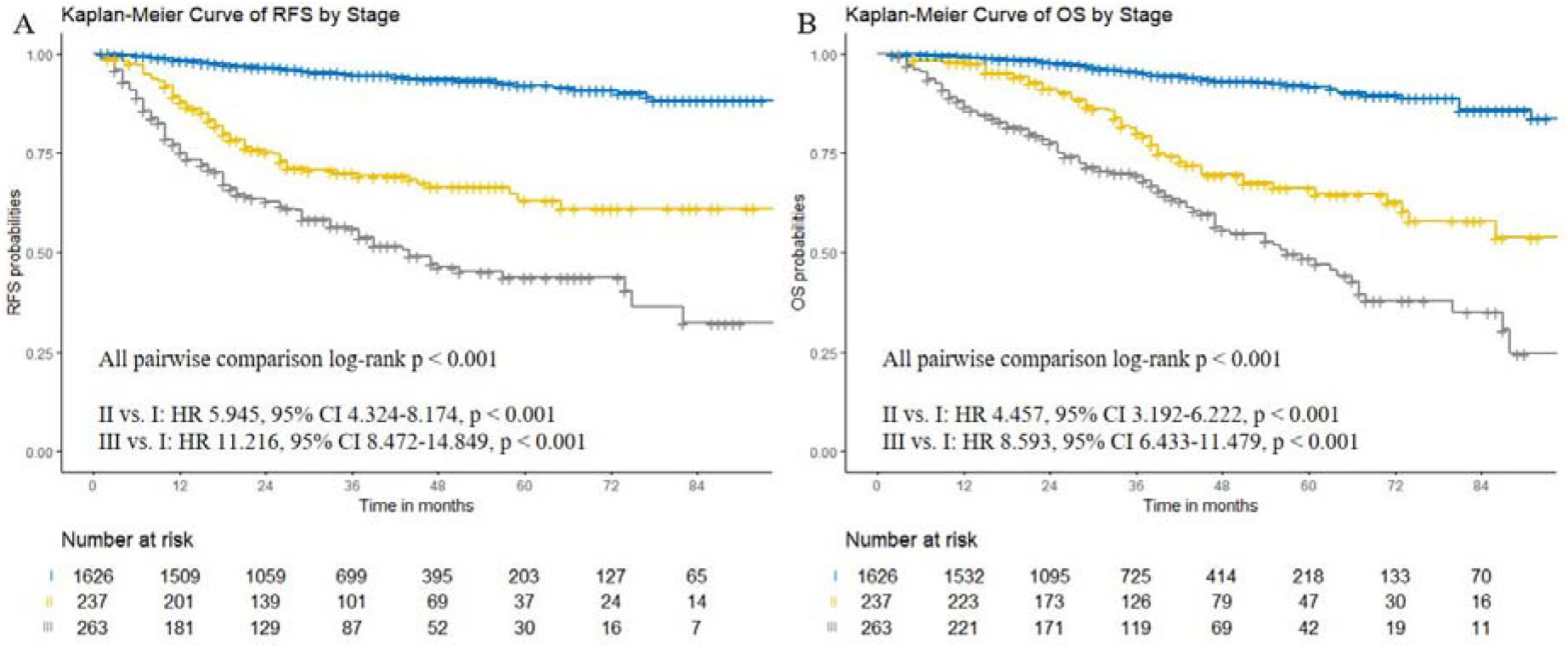
Kaplan-Meier curves showing survival by stage in entire cohort. (A) recurrence-free survival, (B) overall survival. Comparisons were conducted using log-rank test.

Subgroup analysis stratified by stage was then performed to assess whether AI-detected TNN was an independent prognostic factor for patients in each stage. We found that TNN was not significantly associated with survival for patients with stage I (RFS: HR 1.010, 95% CI 0.998-1.022, p = 0.102; OS: HR 1.003, 95% CI 0.989-1.017, p = 0.689) and stage II disease (RFS: HR 1.000, 95% CI 0.988-1.013, p = 0.973; OS: HR 1.000, 95% CI 0.989-1.012, p = 0.965). However, in stage III cohort, fewer TNN was independently associated with improved survival in multivariate analyses (RFS: HR 1.012, 95% CI 1.002-1.022, p = 0.021; OS: HR 1.013, 95% CI 1.002-1.025, p = 0.021) (Table 3 and Table 4).

**Table 3.**
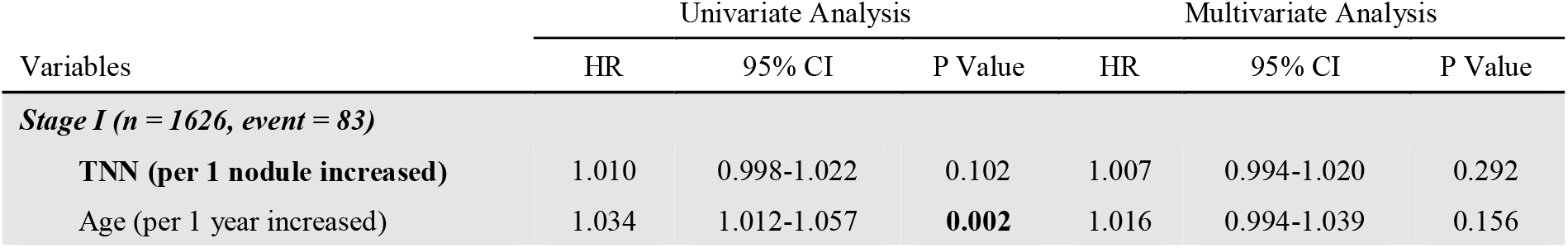
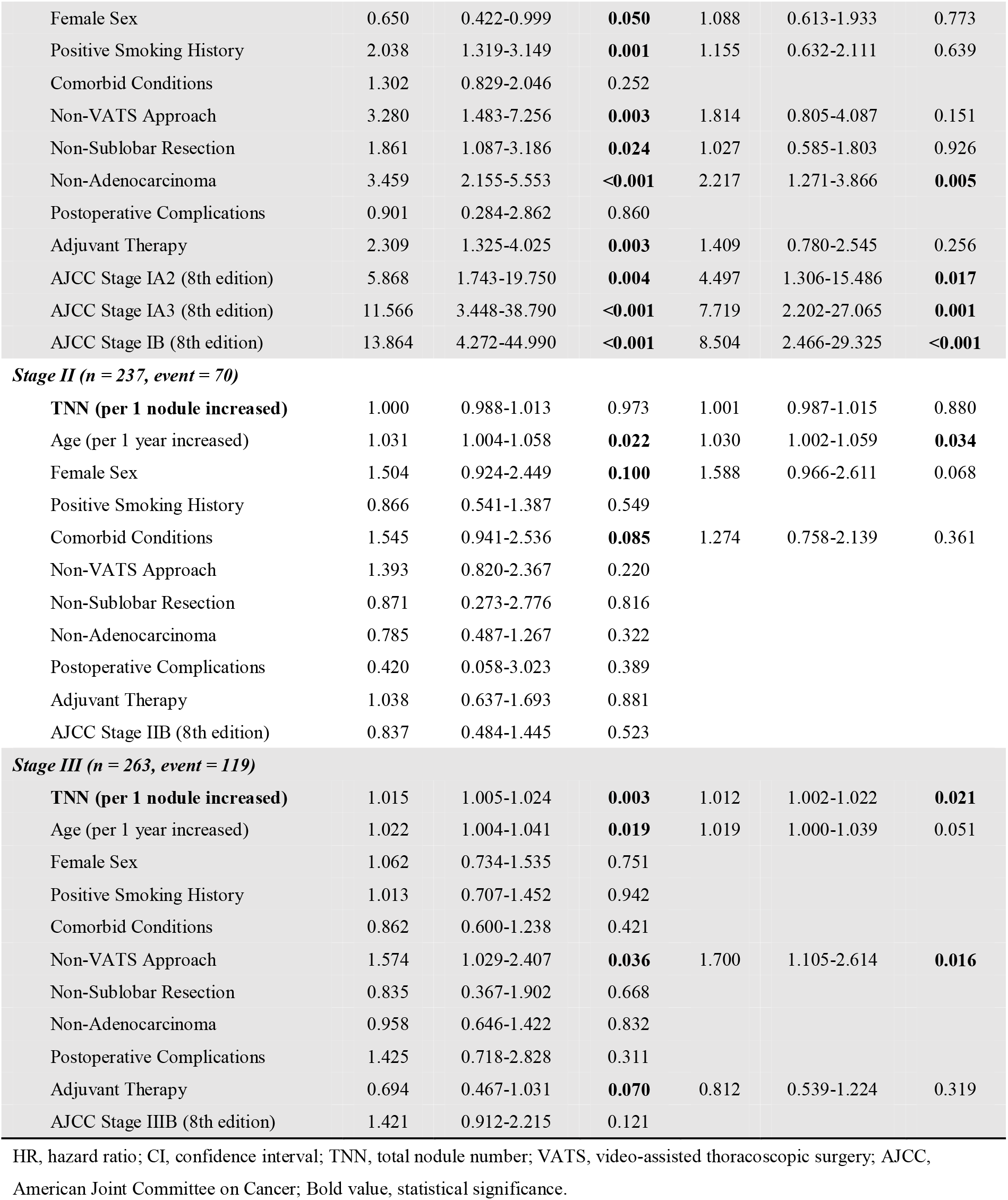
Univariate and Multivariate Analyses of Recurrence-Free Survival (RFS) Stratified by Stage.

| Variables | Univariate Analysis |  |  | Multivariate Analysis |  |  |
| --- | --- | --- | --- | --- | --- | --- |
|  | HR | 95% CI | P Value | HR | 95% CI | P Value |
| **Stage I (n = 1626, event = 83)** |  |  |  |  |  |  |
| **TNN (per 1 nodule increased)** | 1.010 | 0.998-1.022 | 0.102 | 1.007 | 0.994-1.020 | 0.292 |
| Age (per 1 year increased) | 1.034 | 1.012-1.057 | **0.002** | 1.016 | 0.994-1.039 | 0.156 |
| Female Sex | 0.650 | 0.422-0.999 | **0.050** | 1.088 | 0.613-1.933 | 0.773 |
| Positive Smoking History | 2.038 | 1.319-3.149 | **0.001** | 1.155 | 0.632-2.111 | 0.639 |
| Comorbid Conditions | 1.302 | 0.829-2.046 | 0.252 |  |  |  |
| Non-VATS Approach | 3.280 | 1.483-7.256 | **0.003** | 1.814 | 0.805-4.087 | 0.151 |
| Non-Sublobar Resection | 1.861 | 1.087-3.186 | **0.024** | 1.027 | 0.585-1.803 | 0.926 |
| Non-Adenocarcinoma | 3.459 | 2.155-5.553 | **<0.001** | 2.217 | 1.271-3.866 | **0.005** |
| Postoperative Complications | 0.901 | 0.284-2.862 | 0.860 |  |  |  |
| Adjuvant Therapy | 2.309 | 1.325-4.025 | **0.003** | 1.409 | 0.780-2.545 | 0.256 |
| AJCC Stage IA2 (8th edition) | 5.868 | 1.743-19.750 | **0.004** | 4.497 | 1.306-15.486 | **0.017** |
| AJCC Stage IA3 (8th edition) | 11.566 | 3.448-38.790 | **<0.001** | 7.719 | 2.202-27.065 | **0.001** |
| AJCC Stage IB (8th edition) | 13.864 | 4.272-44.990 | **<0.001** | 8.504 | 2.466-29.325 | **<0.001** |
| **Stage II (n = 237, event = 70)** |  |  |  |  |  |  |
| **TNN (per 1 nodule increased)** | 1.000 | 0.988-1.013 | 0.973 | 1.001 | 0.987-1.015 | 0.880 |
| Age (per 1 year increased) | 1.031 | 1.004-1.058 | **0.022** | 1.030 | 1.002-1.059 | **0.034** |
| Female Sex | 1.504 | 0.924-2.449 | **0.100** | 1.588 | 0.966-2.611 | 0.068 |
| Positive Smoking History | 0.866 | 0.541-1.387 | 0.549 |  |  |  |
| Comorbid Conditions | 1.545 | 0.941-2.536 | **0.085** | 1.274 | 0.758-2.139 | 0.361 |
| Non-VATS Approach | 1.393 | 0.820-2.367 | 0.220 |  |  |  |
| Non-Sublobar Resection | 0.871 | 0.273-2.776 | 0.816 |  |  |  |
| Non-Adenocarcinoma | 0.785 | 0.487-1.267 | 0.322 |  |  |  |
| Postoperative Complications | 0.420 | 0.058-3.023 | 0.389 |  |  |  |
| Adjuvant Therapy | 1.038 | 0.637-1.693 | 0.881 |  |  |  |
| AJCC Stage IIB (8th edition) | 0.837 | 0.484-1.445 | 0.523 |  |  |  |
| **Stage III (n = 263, event = 119)** |  |  |  |  |  |  |
| **TNN (per 1 nodule increased)** | 1.015 | 1.005-1.024 | **0.003** | 1.012 | 1.002-1.022 | **0.021** |
| Age (per 1 year increased) | 1.022 | 1.004-1.041 | **0.019** | 1.019 | 1.000-1.039 | 0.051 |
| Female Sex | 1.062 | 0.734-1.535 | 0.751 |  |  |  |
| Positive Smoking History | 1.013 | 0.707-1.452 | 0.942 |  |  |  |
| Comorbid Conditions | 0.862 | 0.600-1.238 | 0.421 |  |  |  |
| Non-VATS Approach | 1.574 | 1.029-2.407 | **0.036** | 1.700 | 1.105-2.614 | **0.016** |
| Non-Sublobar Resection | 0.835 | 0.367-1.902 | 0.668 |  |  |  |
| Non-Adenocarcinoma | 0.958 | 0.646-1.422 | 0.832 |  |  |  |
| Postoperative Complications | 1.425 | 0.718-2.828 | 0.311 |  |  |  |
| Adjuvant Therapy | 0.694 | 0.467-1.031 | **0.070** | 0.812 | 0.539-1.224 | 0.319 |
| AJCC Stage IIIB (8th edition) | 1.421 | 0.912-2.215 | 0.121 |  |  |  |

HR, hazard ratio; CI, confidence interval; TNN, total nodule number; VATS, video-assisted thoracoscopic surgery; AJCC, American Joint Committee on Cancer; Bold value, statistical significance.

**Table 4.**
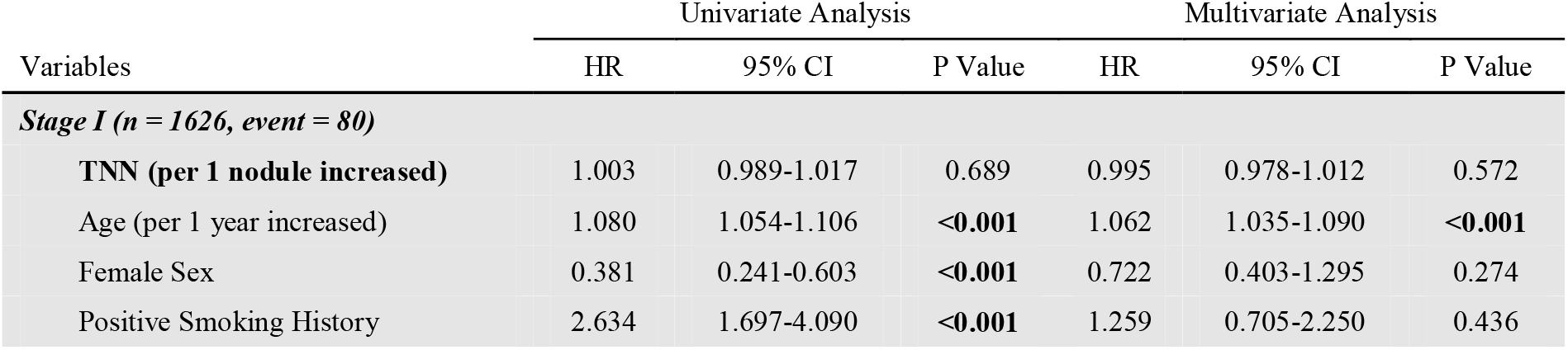
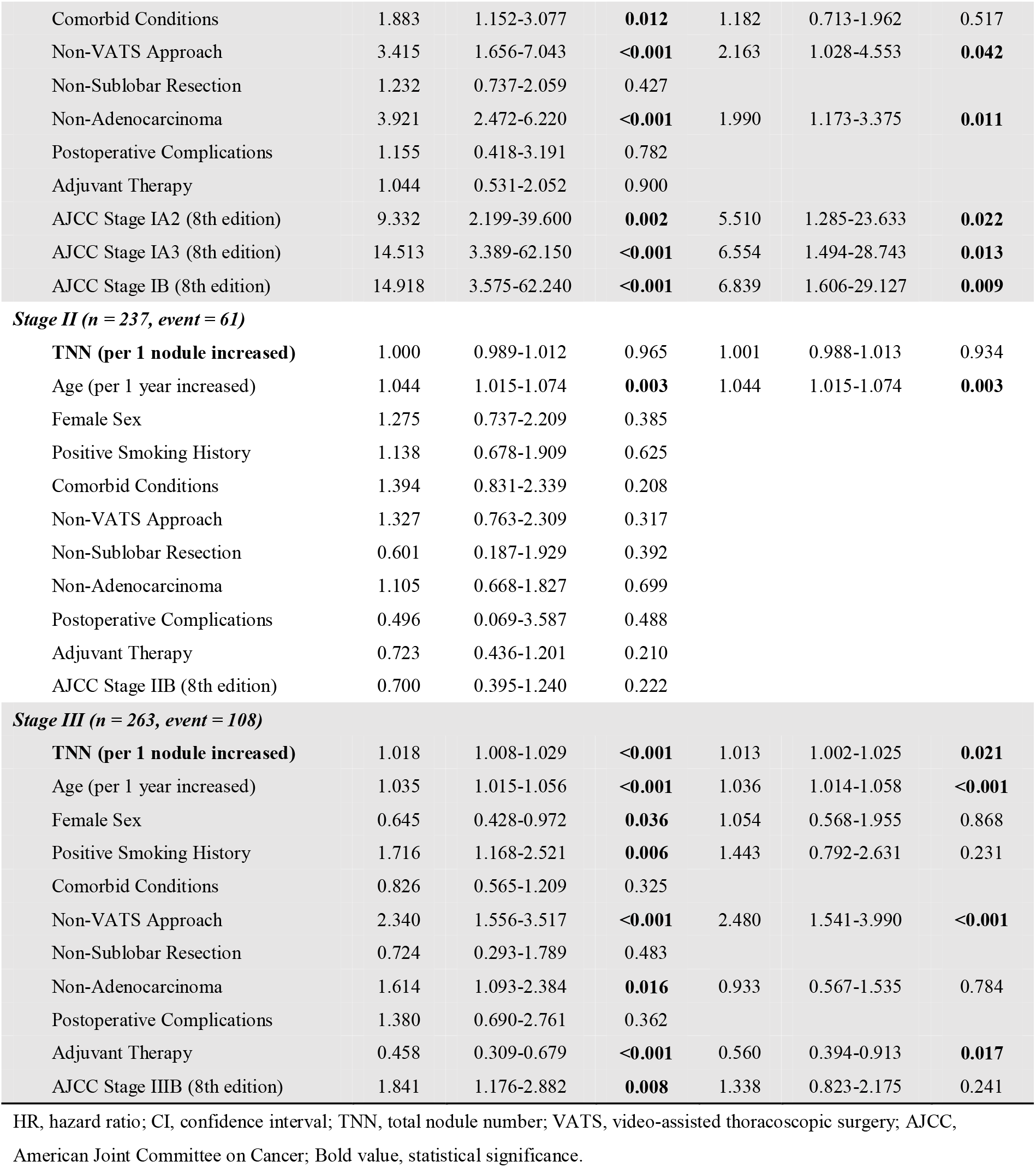
Univariate and Multivariate Analyses of Overall Survival (OS) Stratified by Stage.

| Variables | Univariate Analysis |  |  | Multivariate Analysis |  |  |
| --- | --- | --- | --- | --- | --- | --- |
|  | HR | 95% CI | P Value | HR | 95% CI | P Value |
| **Stage I (n = 1626, event = 80)** |  |  |  |  |  |  |
| **TNN (per 1 nodule increased)** | 1.003 | 0.989-1.017 | 0.689 | 0.995 | 0.978-1.012 | 0.572 |
| Age (per 1 year increased) | 1.080 | 1.054-1.106 | **<0.001** | 1.062 | 1.035-1.090 | **<0.001** |
| Female Sex | 0.381 | 0.241-0.603 | **<0.001** | 0.722 | 0.403-1.295 | 0.274 |
| Positive Smoking History | 2.634 | 1.697-4.090 | **<0.001** | 1.259 | 0.705-2.250 | 0.436 |
| Comorbid Conditions | 1.883 | 1.152-3.077 | **0.012** | 1.182 | 0.713-1.962 | 0.517 |
| Non-VATS Approach | 3.415 | 1.656-7.043 | **<0.001** | 2.163 | 1.028-4.553 | **0.042** |
| Non-Sublobar Resection | 1.232 | 0.737-2.059 | 0.427 |  |  |  |
| Non-Adenocarcinoma | 3.921 | 2.472-6.220 | **<0.001** | 1.990 | 1.173-3.375 | **0.011** |
| Postoperative Complications | 1.155 | 0.418-3.191 | 0.782 |  |  |  |
| Adjuvant Therapy | 1.044 | 0.531-2.052 | 0.900 |  |  |  |
| AJCC Stage IA2 (8th edition) | 9.332 | 2.199-39.600 | **0.002** | 5.510 | 1.285-23.633 | **0.022** |
| AJCC Stage IA3 (8th edition) | 14.513 | 3.389-62.150 | **<0.001** | 6.554 | 1.494-28.743 | **0.013** |
| AJCC Stage IB (8th edition) | 14.918 | 3.575-62.240 | **<0.001** | 6.839 | 1.606-29.127 | **0.009** |
| **Stage II (n = 237, event = 61)** |  |  |  |  |  |  |
| **TNN (per 1 nodule increased)** | 1.000 | 0.989-1.012 | 0.965 | 1.001 | 0.988-1.013 | 0.934 |
| Age (per 1 year increased) | 1.044 | 1.015-1.074 | **0.003** | 1.044 | 1.015-1.074 | **0.003** |
| Female Sex | 1.275 | 0.737-2.209 | 0.385 |  |  |  |
| Positive Smoking History | 1.138 | 0.678-1.909 | 0.625 |  |  |  |
| Comorbid Conditions | 1.394 | 0.831-2.339 | 0.208 |  |  |  |
| Non-VATS Approach | 1.327 | 0.763-2.309 | 0.317 |  |  |  |
| Non-Sublobar Resection | 0.601 | 0.187-1.929 | 0.392 |  |  |  |
| Non-Adenocarcinoma | 1.105 | 0.668-1.827 | 0.699 |  |  |  |
| Postoperative Complications | 0.496 | 0.069-3.587 | 0.488 |  |  |  |
| Adjuvant Therapy | 0.723 | 0.436-1.201 | 0.210 |  |  |  |
| AJCC Stage IIB (8th edition) | 0.700 | 0.395-1.240 | 0.222 |  |  |  |
| **Stage III (n = 263, event = 108)** |  |  |  |  |  |  |
| **TNN (per 1 nodule increased)** | 1.018 | 1.008-1.029 | **<0.001** | 1.013 | 1.002-1.025 | **0.021** |
| Age (per 1 year increased) | 1.035 | 1.015-1.056 | **<0.001** | 1.036 | 1.014-1.058 | **<0.001** |
| Female Sex | 0.645 | 0.428-0.972 | **0.036** | 1.054 | 0.568-1.955 | 0.868 |
| Positive Smoking History | 1.716 | 1.168-2.521 | **0.006** | 1.443 | 0.792-2.631 | 0.231 |
| Comorbid Conditions | 0.826 | 0.565-1.209 | 0.325 |  |  |  |
| Non-VATS Approach | 2.340 | 1.556-3.517 | **<0.001** | 2.480 | 1.541-3.990 | **<0.001** |
| Non-Sublobar Resection | 0.724 | 0.293-1.789 | 0.483 |  |  |  |
| Non-Adenocarcinoma | 1.614 | 1.093-2.384 | **0.016** | 0.933 | 0.567-1.535 | 0.784 |
| Postoperative Complications | 1.380 | 0.690-2.761 | 0.362 |  |  |  |
| Adjuvant Therapy | 0.458 | 0.309-0.679 | **<0.001** | 0.560 | 0.394-0.913 | **0.017** |
| AJCC Stage IIIB (8th edition) | 1.841 | 1.176-2.882 | **0.008** | 1.338 | 0.823-2.175 | 0.241 |

HR, hazard ratio; CI, confidence interval; TNN, total nodule number; VATS, video-assisted thoracoscopic surgery; AJCC, American Joint Committee on Cancer; Bold value, statistical significance.

### Exploratory Analyses in Stage III Cohort

To further evaluate the prognostic effect of AI-detected TNN, we used maximally selected log-rank statistics to dichotomize patients into lower- and higher-TNN groups. The optimal cutoff value of 8 was selected (Supplementary Figure 2). When compared with patients with higher TNN (> 8), those with lower TNN (≤ 8) had significantly improved OS (log-rank p < 0.001, Figure 4A). Lower TNN was also an independent favorable predictor for OS in multivariate analysis (HR 2.348, 95% CI 1.351-4.082, p = 0.002).

**Figure 4.**
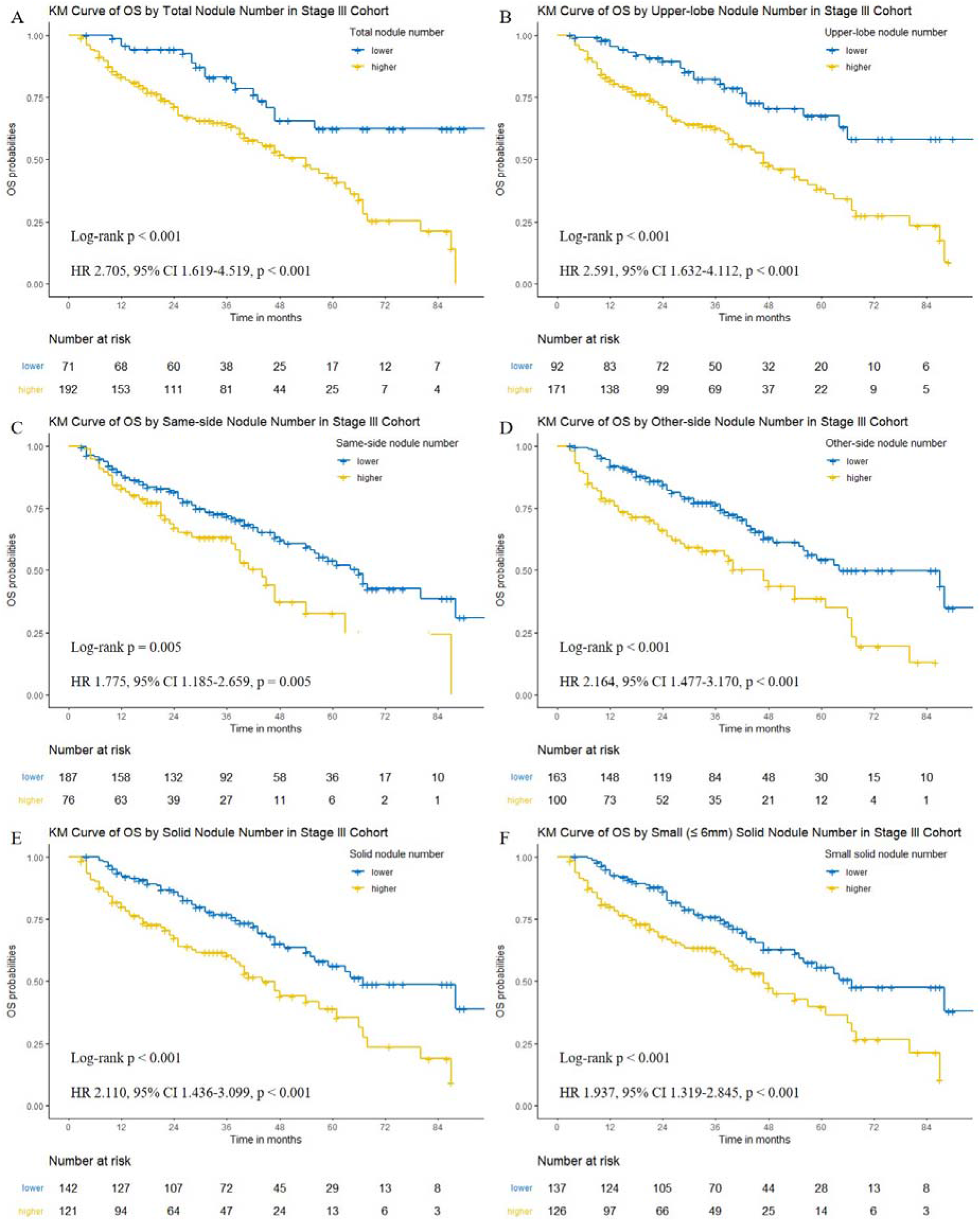
Kaplan-Meier curves showing overall survival by AI-detected nodule number in stage III cohort. (A) TNN, (B) upper-lobe nodule number, (C) same-side nodule number, (D) other-side nodule number, (E) solid nodule number, (F) small (≤ 6mm) solid nodule number. Comparisons were conducted using log-rank test. TNN, total nodule number.

To assess which of the components were associated with survival, we classified AI-detected nodules into different categories. When analyzed as continuous variables, the numbers of upper-lobe nodule (HR 1.028, 95% CI 1.008-1.049, p = 0.006), same-side nodule (HR 1.032, 95% CI 1.001-1.064, p = 0.046), other-side nodule (HR 1.020, 95% CI 1.001-1.039, p = 0.040), solid nodule (HR 1.020, 95% CI 1.004-1.036, p = 0.012), and even solid nodule at small size (≤ 6mm) (HR 1.027, 95% CI 1.007-1.047, p = 0.008) were independently associated with OS in multivariate analyses; however, none of the numbers of middle/lower-lobe nodule (HR 1.016, 95% CI 0.994-1.039, p = 0.153), same-lobe nodule (HR 1.021, 95% CI 0.986-1.056, p = 0.246), m-GGN (HR 1.104, 95% CI 0.885-1.376, p = 0.381), p-GGN (HR 1.015, 95% CI 0.976-1.056, p = 0.462), calcific nodule (HR 1.021, 95% CI 0.975-1.068, p = 0.384), and perifissural nodule (HR 1.007, 95% CI 0.792-1.279, p = 0.957) were significantly associated with survival.

The five above-mentioned nodule numbers, which were independent prognostic factors for OS as continuous variables, were set as binary variables according to their optimal cutoff values. Similarly, compared with patients with higher nodule numbers, those with lower nodule numbers had significantly improved OS (Figure 4B to F). In addition, the numbers of upper-lobe nodule (HR 2.532, 95% CI 1.567-4.091, p < 0.001), other-side nodule (HR 1.957, 95% CI 1.322-2.898, p < 0.001), solid nodule (HR 1.851, 95% CI 1.241-2.761, p = 0.003), and solid nodule at small size (≤ 6mm) (HR 1.862, 95% CI 1.248-2.779, p = 0.002) were still independent prognostic factors for OS in multivariate analyses.

Finally, to evaluate which of the components contributed most to prognosis, LASSO Cox regression model incorporating both clinicopathologic features and all categories of AI-detected nodule numbers (as continuous variables) was built (Supplementary Figure 3). Seven features with nonzero coefficient were as follows: age (0.021), smoking history (0.106), surgical approach (0.669), adjuvant therapy status (−0.389), IIIA/IIIB classification (0.095), upper-lobe nodule number (0.014), and small (≤ 6mm) solid nodule number (0.008). Thus, the number of upper-lobe nodule, followed by solid nodule at small size, were individual features that contributed most to the model and corelated best with OS among all categories of AI-detected nodule numbers.

### Survival Tree Analyses

A tree-based model incorporating AI-detected TNN and the 8^th^ edition of AJCC prognostic group was constructed based on the best split of OS in the entire cohort (Figure 5A). We found that discrimination of survival curves of sub-stages was unsatisfactory with the current staging system in our study, especially in sub-stages of IA2 to IB (IA2 vs. IA3: log-rank p = 0.177; IA3 vs. IB: log-rank p = 0.778) and IIA to IIB (log-rank p = 0.236). Moreover, in stage III cohort, rather than using the traditional IIIA and IIIB classification, the model grouped OS by AI-detected TNN (lower vs. higher: log-rank p < 0.001), since it showed superior discrimination of survival. Accordingly, Kaplan-Meier curves of OS by the tree-based grouping scheme were shown in Figure 5B.

**Figure 5.**
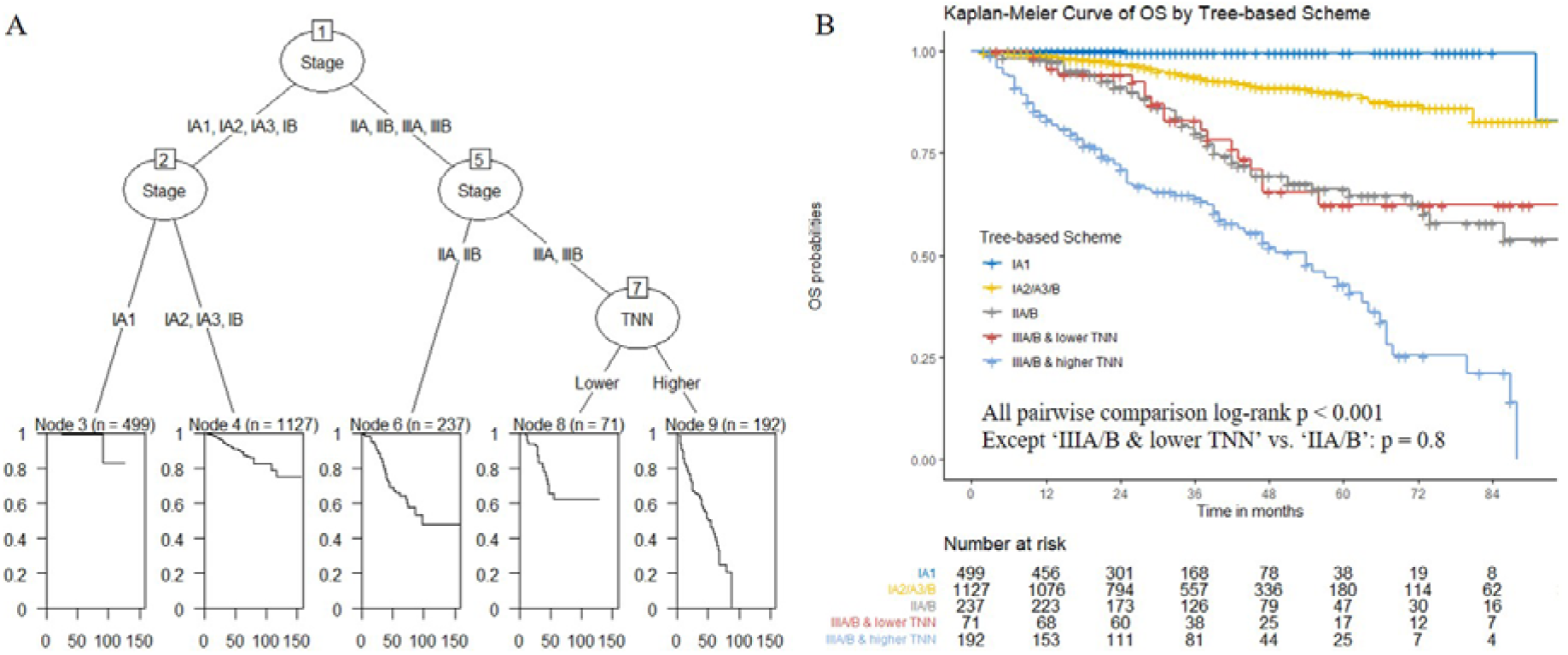
Survival tree analysis. (A) Recursive partitioning-generated survival tree based on the best split of overall survival using AI-detected TNN and the 8^th^ edition of AJCC stage. Both TNN and stage were modeled as categorical variables. (B) Kaplan-Meier curves showing overall survival by tree-based scheme in entire cohort. Comparisons were conducted using log-rank test. AJCC, American Joint Committee on Cancer; TNN, total nodule number.

### Treatment Failure Analyses

To evaluate the potential relationship between AI-detected TNN and the tumor recurrence pattern, we further divided stage III cohort into two groups according to the first disease progression site. Among all 263 patients, 60 had local recurrent disease, 40 had distant metastatic disease, and 19 had progressive disease without specified pattern. Compared with distant metastasis group (median: 17 [IQR 10.75-23.25]), patients with local recurrent disease had lower AI-detected TNN (median: 14 [IQR 7.75-18.25]). However, the difference between these two groups was not statistically significant (Wilcoxon rank-sum p = 0.077, Supplementary Figure 4).

## DISCUSSION

The widespread application of artificial intelligence algorithm in PN detection is reshaping our knowledge on this topic. The number of patients with tens or even hundreds of PNs is rapidly growing, while the interpretation of these lesions and its impact on surgical decision making are complicated and yet underrepresented. As the number of nodules grows, accurate diagnosis for every single nodule becomes labor-intensive and statistically challenging. As an alternative, we hypothesized that TNN measured by deep-learning algorithm may serve as a surrogate indicator of the probability of malignancy and metastasis in locally advanced NSCLC. Such hypothesis is proved preliminarily by our result that TNN is an independent prognostic factor in stage III lung cancer.

The accurate measurement of TNN is highly challenging. First, the definition of PN varies across radiologists and surgeons due to their different purposes: some may only report guideline mandated PNs in order not to provoke panic of patients, while others may report as many as one can detect for more accurate surgery planning. Unfortunately, both standards are rather subjective and less repeatable. Second, the accuracy and robustness of a single radiologist or surgeon is limited. The sensitivity of PN detection by a single radiologist is around 77% and can be increased to 90% with a concurrent radiologist’s help [28]. However, such method is time-consuming and still subjected to human error.

The emerging of deep learning-based AI algorithm ensured the objectiveness and robustness of PN detection and thus the measurement of TNN. Mature algorithms have reached a diagnosis sensitivity of 85-100% [29-31]. The best-performing deep learning algorithm in LUNA16 challenge, which based on the LIDC-IDRI dataset, exhibited an excellent sensitivity of over 95% at fewer than 1.0 false positives per scan [32]. The algorithm (InferRead™ CT Lung, InferVision) in this study is trained using over 350 thousand chest CTs labeled by radiologists [33]. In real-world, the performance of this model has reached AUC at 0.89 in PN detection and can significantly improve the performance assisting radiologists [33-35]. Our result showed median TNN at 12 per patient, much higher than the median of 2 per patient reported in the malignant cohort of NELSON study [26]. Such difference may, on one hand, because of the difference in CT radiation dosage, while on the other hand, reflect the difference in diagnosis preference and consistency between AI and human radiologists.

From the clinical aspect, our results suggested that TNN may be a visualized representation of tumor burden in stage III NSCLC patients. Different from the result of NELSON study that shows higher nodule count is in favor of benign diagnosis [26], our study focused on more advanced NSCLC patients instead of high-risk screening population. Past evidences vaguely showed that, with confirmed histology, extensive nodal or systemic metastasis are relative arguments for multiple PNs to be IPM [36], suggesting that high TNN may relate to higher pretest probability of IPM. Our result further supported this speculation by revealing the survival advantage of lower TNN group comparing to higher TNN. Such advantage existed when analyzing TNN as either a continuous variable or a binary variable, which strengthened the argument.

It is worth notice that the major impact on survival was caused by the number of solid nodules but not GGNs. For GGN components, the IASLC guideline suggested that the prognosis of multifocal GGNs is similar to single MIA or AIS [37], while others indicated that there are metastatic GGNs on molecular level [38]. In our study, concurrent multiple GGNs in all three stages did not increase HR, indicating that concurrent multiple GGNs in invasive lung cancer possessed the same biological behavior as in multifocal GGN cases. For solid components, most of the nodules were ≤ 6mm and radiologically benign, with round shape, no speculation, no lobulation. But the growth of a few unresected nodules suggested their malignancy (Supplementary Figure 5). It showed that the diagnosis using traditional radiological characteristics for multiple PNs in stage III NSCLC patients is less reliable. The treatment failure pattern analysis showed that higher TNN was related to distant metastasis (without statistical significance due to small sample size) indicating that TNN was not only an indicator of IPM, but also a visual representation of systematic tumor burden.

From a surgeon’s perspective, the impact of PNs on surgical planning is dramatic. Convincing a patient to accept unresected GGNs after surgery is difficult even if with the guideline’s support. A sublobar resection of a GGN may turn into lobectomy due to multiple GGNs detected by AI, while lobectomy may also be altered into sublobar resection due to bilateral nodules clinically diagnosed as separate primary lung cancer. However, no evidence showed the validity of such approach. Our study provided the first proof of concept that TNN, driven by deep learning algorithm, should be considered as a mandatory test before surgery planning. It would be reasonable for surgeons to be more aggressive in resection of solid nodules instead of GGNs. Moreover, neoadjuvant therapy should be considered for stage III patients with higher TNN for better PN evaluation since empirical diagnosis may not be reliable.

Some may argue that PET-CT is a valid method in differentiating MPLC and IPM before surgery, but the partial-volume effect of PET-CT prevented it from a proper diagnostic performance for solid nodules less than 8mm, which represented 85.6% of the solid nodules in our study [23, 39, 40]. Moreover, PET-CT is relatively expensive for most under-development countries and is not affordable by every patient.

As a retrospective study, our results need validation before clinical application, however, no public databases provide sufficient data, thus a prospective validation is needed and may be time-consuming. The AI algorithm needs optimization to further reduce false positive rate, and the performance in the detection of peri-vascular nodule still needs improvement. Currently, there is a technological barrier in the alignment of pre- and post-operative PNs on chest CT. Given time, we may be able to analyze the growth speed of PN and other information for better prognostic modeling.

To our knowledge, this study is the first to identify that TNN measured by deep-learning algorithm is an independent prognostic factor in stage III lung cancer. Our results suggested a potentially critical clinical application of AI as a mandatory examination for surgery decision. The current cut-off point of TNN is still preliminary but shows great potential and appeals to future validation.

## Data Availability

The data used to support the findings of this study are available from the corresponding author upon request.

## AUTHOR CONTRIBUTIONS

XC took full responsibility for the content of the manuscript. QQ, and ZS had full access to all the data in the study and took responsibility for the integrity of the data and the accuracy of the data analysis. DW, JS, WT, XL, TL, NH, and FY contributed substantially to the study design, data analysis and interpretation.

## ACKNOWLEDGEMENTS

We would like to thank Yuqing Huang, Xianjun Min, and Guotian Pei from Beijing Haidian Hospital for sharing their thoughts on this work.

## Abbreviations list

AI: artificial intelligence
CAD: computer aided detection/diagnosis
CNN: convolutional neural network
GGN: ground-glass nodule
HR: hazard ratio
IPM: intrapulmonary metastasis
LDCT: low-dose computed tomography
MPLC: multiple primary lung cancer
NSCLC: non-small cell lung cancer
OS: overall survival
PET: positron emission tomography
PN: pulmonary nodule
RFS: recurrence-free survival
TNN: total nodule number

## REFERENCES

1. Bray F, Ferlay J, Soerjomataram I, Siegel RL, Torre LA, Jemal A. Global cancer statistics 2018: GLOBOCAN estimates of incidence and mortality worldwide for 36 cancers in 185 countries. CA Cancer J Clin. 2018;68(6):394–424.

2. Aberle DR, Adams AM, Berg CD, Black WC, Clapp JD, Fagerstrom RM, et al. Reduced lung-cancer mortality with low-dose computed tomographic screening. N Engl J Med. 2011;365(5):395–409.

3. Detterbeck FC, Mazzone PJ, Naidich DP, Bach PB. Screening for lung cancer: Diagnosis and management of lung cancer, 3rd ed: American College of Chest Physicians evidence-based clinical practice guidelines. Chest. 2013;143(5 Suppl):e78S–e92S.

4. Ruparel M, Quaife SL, Navani N, Wardle J, Janes SM, Baldwin DR. Pulmonary nodules and CT screening: the past, present and future. Thorax. 2016;71(4):367–75.

5. Smith RA, Andrews KS, Brooks D, Fedewa SA, Manassaram-Baptiste D, Saslow D, et al. Cancer screening in the United States, 2017: A review of current American Cancer Society guidelines and current issues in cancer screening. CA Cancer J Clin. 2017;67(2):100–21.

6. Wood DE, Kazerooni EA, Baum SL, Eapen GA, Ettinger DS, Hou L, et al. Lung Cancer Screening, Version 3.2018, NCCN Clinical Practice Guidelines in Oncology. J Natl Compr Canc Netw. 2018;16(4):412–41.

7. de Koning HJ, van der Aalst CM, de Jong PA, Scholten ET, Nackaerts K, Heuvelmans MA, et al. Reduced Lung-Cancer Mortality with Volume CT Screening in a Randomized Trial. N Engl J Med. 2020;382(6):503–13.

8. Gould MK, Tang T, Liu IL, Lee J, Zheng C, Danforth KN, et al. Recent Trends in the Identification of Incidental Pulmonary Nodules. Am J Respir Crit Care Med. 2015;192(10):1208–14.

9. Erickson BJ, Korfiatis P, Akkus Z, Kline TL. Machine Learning for Medical Imaging. Radiographics. 2017;37(2):505–15.

10. Armato SG, Altman MB, L. Rivière PJ. Automated detection of lung nodules in CT scans: effect of image reconstruction algorithm. Med Phys. 2003;30(3):461–72.

11. Hwang EJ, Park CM. Clinical Implementation of Deep Learning in Thoracic Radiology: Potential Applications and Challenges. Korean J Radiol. 2020;21(5):511–25.

12. Ather S, Kadir T, Gleeson F. Artificial intelligence and radiomics in pulmonary nodule management: current status and future applications. Clin Radiol. 2020;75(1):13–9.

13. Murphy A, Skalski M, Gaillard F. The utilisation of convolutional neural networks in detecting pulmonary nodules: a review. Br J Radiol. 2018;91(1090):20180028.

14. Hua KL, Hsu CH, Hidayati SC, Cheng WH, Chen YJ. Computer-aided classification of lung nodules on computed tomography images via deep learning technique. Onco Targets Ther. 2015;8:2015–22.

15. Cheng JZ, Ni D, Chou YH, Qin J, Tiu CM, Chang YC, et al. Computer-Aided Diagnosis with Deep Learning Architecture: Applications to Breast Lesions in US Images and Pulmonary Nodules in CT Scans. Sci Rep. 2016;6:24454.

16. Li W, Cao P, Zhao D, Wang J. Pulmonary Nodule Classification with Deep Convolutional Neural Networks on Computed Tomography Images. Comput Math Methods Med. 2016;2016:6215085.

17. Liu S, Xie Y, Jirapatnakul A, Reeves AP. Pulmonary nodule classification in lung cancer screening with three-dimensional convolutional neural networks. J Med Imaging (Bellingham). 2017;4(4):041308.

18. da Silva Glf, Valente TLA, Silva AC, de Paiva AC, Gattass M. Convolutional neural network-based PSO for lung nodule false positive reduction on CT images. Comput Methods Programs Biomed. 2018;162:109–18.

19. Xie Y, Xia Y, Zhang J, Song Y, Feng D, Fulham M, et al. Knowledge-based Collaborative Deep Learning for Benign-Malignant Lung Nodule Classification on Chest CT. IEEE Trans Med Imaging. 2019;38(4):991–1004.

20. McWilliams A, Tammemagi MC, Mayo JR, Roberts H, Liu G, Soghrati K, et al. Probability of cancer in pulmonary nodules detected on first screening CT. N Engl J Med. 2013;369(10):910–9.

21. Horeweg N, van Rosmalen J, Heuvelmans MA, van der Aalst CM, Vliegenthart R, Scholten ET, et al. Lung cancer probability in patients with CT-detected pulmonary nodules: a prespecified analysis of data from the NELSON trial of low-dose CT screening. Lancet Oncol. 2014;15(12):1332–41.

22. Walter JE, Heuvelmans MA, de Jong PA, Vliegenthart R, van Ooijen Pma, Peters RB, et al. Occurrence and lung cancer probability of new solid nodules at incidence screening with low-dose CT: analysis of data from the randomised, controlled NELSON trial. Lancet Oncol. 2016;17(7):907–16.

23. Gould MK, Donington J, Lynch WR, Mazzone PJ, Midthun DE, Naidich DP, et al. Evaluation of individuals with pulmonary nodules: when is it lung cancer? Diagnosis and management of lung cancer, 3rd ed: American College of Chest Physicians evidence-based clinical practice guidelines. Chest. 2013;143(5 Suppl):e93S–e120S.

24. Callister ME, Baldwin DR, Akram AR, Barnard S, Cane P, Draffan J, et al. British Thoracic Society guidelines for the investigation and management of pulmonary nodules. Thorax. 2015;70 Suppl 2:ii1–ii54.

25. MacMahon H, Naidich DP, Goo JM, Lee KS, Leung ANC, Mayo JR, et al. Guidelines for Management of Incidental Pulmonary Nodules Detected on CT Images: From the Fleischner Society 2017. Radiology. 2017;284(1):228–43.

26. Heuvelmans MA, Walter JE, Peters RB, Bock GH, Yousaf-Khan U, Aalst CMV, et al. Relationship between nodule count and lung cancer probability in baseline CT lung cancer screening: The NELSON study. Lung Cancer. 2017;113:45–50.

27. Walter JE, Heuvelmans MA, de Bock GH, Yousaf-Khan U, Groen HJM, van der Aalst CM, et al. Relationship between the number of new nodules and lung cancer probability in incidence screening rounds of CT lung cancer screening: The NELSON study. Lung Cancer. 2018;125:103–8.

28. Nair A, Screaton NJ, Holemans JA, Jones D, Clements L, Barton B, et al. The impact of trained radiographers as concurrent readers on performance and reading time of experienced radiologists in the UK Lung Cancer Screening (UKLS) trial. Eur Radiol. 2018;28(1):226–34.

29. Setio AA, Ciompi F, Litjens G, Gerke P, Jacobs C, van Riel SJ, et al. Pulmonary Nodule Detection in CT Images: False Positive Reduction Using Multi-View Convolutional Networks. IEEE Trans Med Imaging. 2016;35(5):1160–9.

30. Dou Q, Chen H, Yu L, Qin J, Heng PA. Multilevel Contextual 3-D CNNs for False Positive Reduction in Pulmonary Nodule Detection. IEEE Trans Biomed Eng. 2017;64(7):1558–67.

31. Tajbakhsh N, Suzuki K. Comparing two classes of end-to-end machine-learning models in lung nodule detection and classification: MTANNs vs. CNNs. Pattern Recognition. 2017. p. 476–86.

32. Setio AAA, Traverso A, de Bel T, Berens MSN, Bogaard CVD, Cerello P, et al. Validation, comparison, and combination of algorithms for automatic detection of pulmonary nodules in computed tomography images: The LUNA16 challenge. Med Image Anal. 2017;42:1–13.

33. Wang Y, Yan F, Lu X, Zheng G, Zhang X, Wang C, et al. IILS: Intelligent imaging layout system for automatic imaging report standardization and intra-interdisciplinary clinical workflow optimization. EBioMedicine. 2019;44:162–81.

34. Liu K, Li Q, Ma J, Zhou Z, Sun M, Deng Y, et al. Evaluating a Fully Automated Pulmonary Nodule Detection Approach and Its Impact on Radiologist Performance. Radiology: Artificial Intelligence. 2019.

35. Yang F, Fan J, Tianzhou J, Yang F, Li Y, Liu X, et al. Population-based research of pulmonary subsolid nodule CT screening and artificial intelligence application. Chin J Thorac Cardiovasc Surg. 2020.

36. Detterbeck FC, Nicholson AG, Franklin WA, Marom EM, Travis WD, Girard N, et al. The IASLC Lung Cancer Staging Project: Summary of Proposals for Revisions of the Classification of Lung Cancers with Multiple Pulmonary Sites of Involvement in the Forthcoming Eighth Edition of the TNM Classification. J Thorac Oncol. 2016;11(5):639–50.

37. Detterbeck FC, Marom EM, Arenberg DA, Franklin WA, Nicholson AG, Travis WD, et al. The IASLC Lung Cancer Staging Project: Background Data and Proposals for the Application of TNM Staging Rules to Lung Cancer Presenting as Multiple Nodules with Ground Glass or Lepidic Features or a Pneumonic Type of Involvement in the Forthcoming Eighth Edition of the TNM Classification. J Thorac Oncol. 2016;11(5):666–80.

38. Li R, Li X, Xue R, Yang F, Wang S, Li Y, et al. Early metastasis detected in patients with multifocal pulmonary ground-glass opacities (GGOs). Thorax. 2018;73(3):290–2.

39. Soret M, Bacharach SL, Buvat I. Partial-volume effect in PET tumor imaging. J Nucl Med. 2007;48(6):932–45.

40. Groheux D, Quere G, Blanc E, Lemarignier C, Vercellino L, de Margerie-Mellon C, et al. FDG PET-CT for solitary pulmonary nodule and lung cancer: Literature review. Diagn Interv Imaging. 2016;97(10):1003–17.

