## Supplemental Tables and Figures for "Total nodule number is an independent prognostic factor in resected stage III non-small cell lung cancer: a deep learning powered study"

Supplementary Material

**Supplementary Table**

| **Supplementary Table. Univariate and Multivariate Survival Analyses of Entire Cohort** | | | | | | |
| --- | --- | --- | --- | --- | --- | --- |
| Variables | Univariate Analysis | | | Multivariate Analysis | | |
|  | HR | 95% CI | P Value | HR | 95% CI | P Value |
| ***Recurrence-free survival (RFS)*** | | | | | | |
| **TNN (per 1 nodule increased)** | 1.008 | 1.001-1.014 | **0.017** | 1.006 | 0.999-1.012 | 0.080 |
| Age (per 1 year increased) | 1.027 | 1.015-1.039 | **<0.001** | 1.033 | 1.019-1.046 | **<0.001** |
| Sex |  |  |  |  |  |  |
| Male | Reference |  |  | Reference |  |  |
| Female | 0.625 | 0.491-0.795 | **<0.001** | 1.249 | 0.884-1.765 | 0.207 |
| Smoking History |  |  |  |  |  |  |
| No | Reference |  |  | Reference |  |  |
| Yes | 1.931 | 1.521-2.451 | **<0.001** | 1.157 | 0.817-1.637 | 0.412 |
| Comorbid Conditions |  |  |  |  |  |  |
| No | Reference |  |  |  |  |  |
| Yes | 1.098 | 0.860-1.401 | 0.453 |  |  |  |
| Surgical Approach |  |  |  |  |  |  |
| VATS | Reference |  |  | Reference |  |  |
| Non-VATS | 1.369 | 2.910-5.313 | **<0.001** | 1.521 | 1.086-2.130 | **0.015** |
| Surgical Procedure |  |  |  |  |  |  |
| Sublobar Resection | Reference |  |  | Reference |  |  |
| Non-Sublobar Resection | 3.601 | 2.401-5.400 | **<0.001** | 1.516 | 0.983-2.338 | 0.060 |
| Histologic Type |  |  |  |  |  |  |
| Adenocarcinoma | Reference |  |  | Reference |  |  |
| Non-Adenocarcinoma | 2.416 | 1.872-3.117 | **<0.001** | 1.070 | 0.773-1.482 | 0.682 |
| Postoperative Complications |  |  |  |  |  |  |
| No | Reference |  |  |  |  |  |
| Yes | 1.225 | 0.701-2.139 | 0.476 |  |  |  |
| Adjuvant Therapy |  |  |  |  |  |  |
| No | Reference |  |  | Reference |  |  |
| Yes | 4.162 | 3.277-5.287 | **<0.001** | 1.330 | 0.987-1.793 | 0.061 |
| AJCC 8th edition, T stage |  |  |  |  |  |  |
| T1 | Reference |  |  | Reference |  |  |
| T2 | 3.449 | 2.602-4.573 | **<0.001** | 1.781 | 1.304-2.431 | **<0.001** |
| T3 | 8.036 | 5.603-11.525 | **<0.001** | 3.568 | 2.386-5.334 | **<0.001** |
| T4 | 10.735 | 6.571-17.537 | **<0.001** | 3.861 | 2.245-6.641 | **<0.001** |
| AJCC 8th edition, N stage |  |  |  |  |  |  |
| N0 | Reference |  |  | Reference |  |  |
| N1 | 4.988 | 3.570-6.969 | **<0.001** | 2.852 | 1.941-4.190 | **<0.001** |
| N2 | 7.501 | 5.760-9.769 | **<0.001** | 4.269 | 3.113-5.855 | **<0.001** |
| ***Overall survival (OS)*** | | | | | | |
| **TNN (per 1 nodule increased)** | 1.006 | 0.999-1.012 | **0.099** | 1.002 | 0.995-1.009 | 0.590 |
| Age (per 1 year increased) | 1.048 | 1.035-1.062 | **<0.001** | 1.057 | 1.042-1.072 | **<0.001** |
| Sex |  |  |  |  |  |  |
| Male | Reference |  |  | Reference |  |  |
| Female | 0.432 | 0.331-0.564 | **<0.001** | 0.961 | 0.665-1.388 | 0.830 |
| Smoking History |  |  |  |  |  |  |
| No | Reference |  |  | Reference |  |  |
| Yes | 2.581 | 2.007-3.319 | **<0.001** | 1.348 | 0.945-1.924 | 0.100 |
| Comorbid Conditions |  |  |  |  |  |  |
| No | Reference |  |  |  |  |  |
| Yes | 1.182 | 0.915-1.527 | 0.200 |  |  |  |
| Surgical Approach |  |  |  |  |  |  |
| VATS | Reference |  |  | Reference |  |  |
| Non-VATS | 4.050 | 3.023-5.426 | **<0.001** | 1.827 | 1.312-2.545 | **<0.001** |
| Surgical Procedure |  |  |  |  |  |  |
| Sublobar Resection | Reference |  |  | Reference |  |  |
| Non-Sublobar Resection | 2.419 | 1.629-3.594 | **<0.001** | 1.089 | 0.711-1.668 | 0.696 |
| Histologic Type |  |  |  |  |  |  |
| Adenocarcinoma | Reference |  |  | Reference |  |  |
| Non-Adenocarcinoma | 2.936 | 2.275-3.790 | **<0.001** | 1.380 | 1.001-1.903 | **0.049** |
| Postoperative Complications |  |  |  |  |  |  |
| No | Reference |  |  |  |  |  |
| Yes | 1.349 | 0.785-2.321 | 0.279 |  |  |  |
| Adjuvant Therapy |  |  |  |  |  |  |
| No | Reference |  |  | Reference |  |  |
| Yes | 2.337 | 1.816-3.007 | **<0.001** | 0.890 | 0.659-1.201 | 0.444 |
| AJCC 8th edition, T stage |  |  |  |  |  |  |
| T1 | Reference |  |  | Reference |  |  |
| T2 | 2.666 | 1.973-3.602 | **<0.001** | 1.455 | 1.045-2.025 | **0.027** |
| T3 | 6.839 | 4.720-9.908 | **<0.001** | 2.900 | 1.922-4.374 | **<0.001** |
| T4 | 11.987 | 7.548-19.037 | **<0.001** | 4.537 | 2.700-7.623 | **<0.001** |
| AJCC 8th edition, N stage |  |  |  |  |  |  |
| N0 | Reference |  |  | Reference |  |  |
| N1 | 3.621 | 2.536-5.171 | **<0.001** | 2.632 | 1.772-3.910 | **<0.001** |
| N2 | 5.490 | 4.162-7.242 | **<0.001** | 4.325 | 3.124-5.988 | **<0.001** |
| HR, hazard ratio; CI, confidence interval; TNN, total nodule number; VATS, video-assisted thoracoscopic surgery; AJCC, American Joint Committee on Cancer; Bold value, statistical significance. | | | | | | |

**Supplementary Figures**

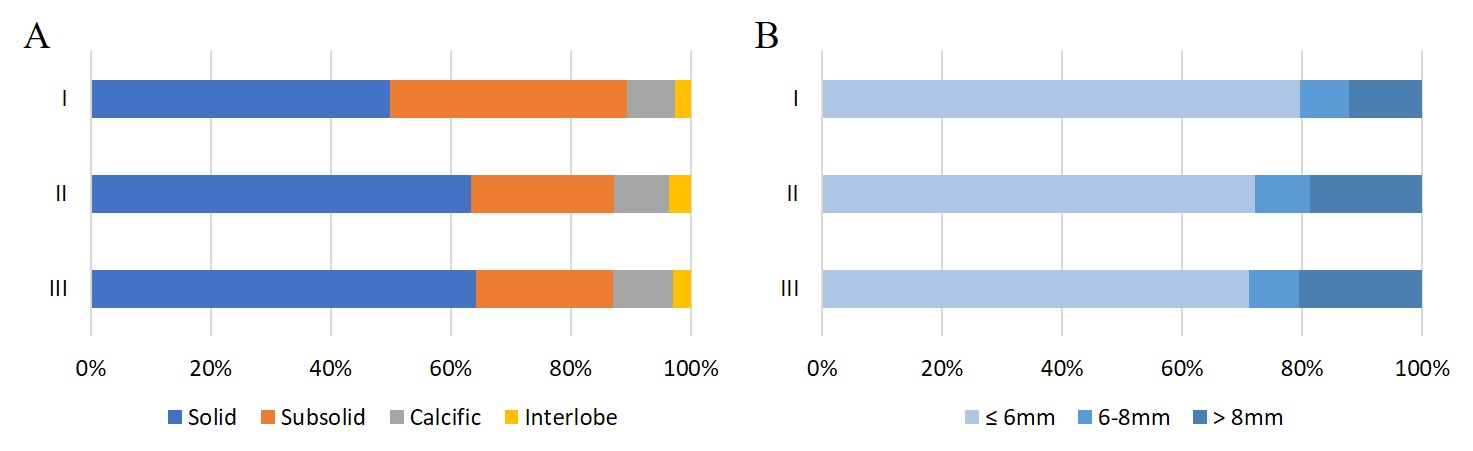

**Supplementary Figure 1. Bar chart showing the percentage of AI-detected nodule categories stratified by pathological stage.** (A) nodule type, (B) solid nodule size.

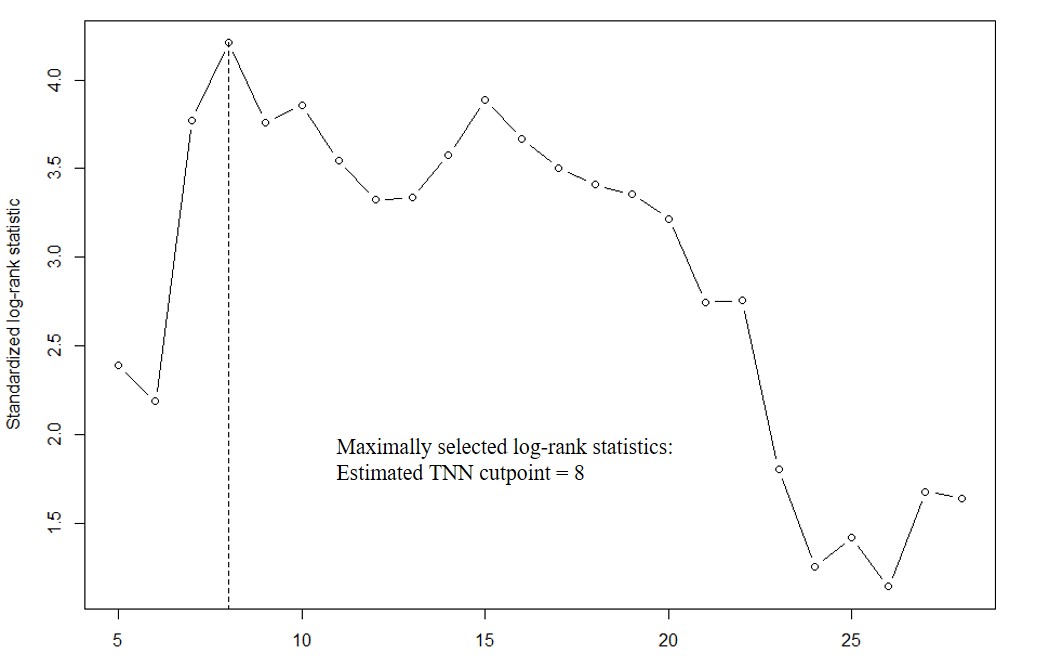

**Supplementary Figure 2. Maximally selected log-rank statistics showing the optimal cutoff value of AI-detected TNN for predicting overall survival.** The optimal cutoff value of 8 was selected. TNN, total nodule number.

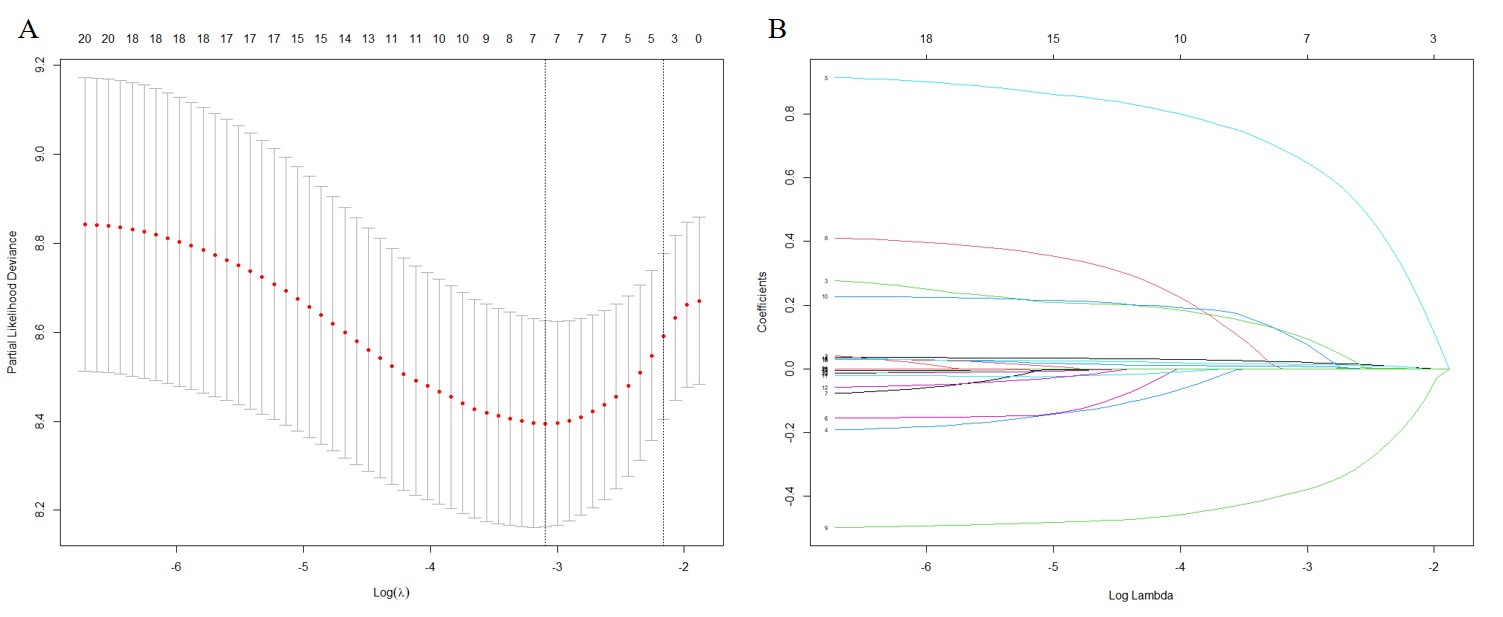

**Supplementary Figure 3. Feature selection with LASSO.** LASSO Cox regression model was built with both clinicopathologic characteristics and all categories of AI-detected nodule numbers (as continuous variables). (A) Red dots indicated average deviance values for each model with a given λ; the dotted vertical lines were drawn at the optimal values by using the minimum mean square error (MSE) criteria. The optimal λ value of 0.0453 was selected. (B) The process of feature selection and resulting features with nonzero coefficients were indicated in the plot. LASSO, least absolute shrinkage and selection operator.

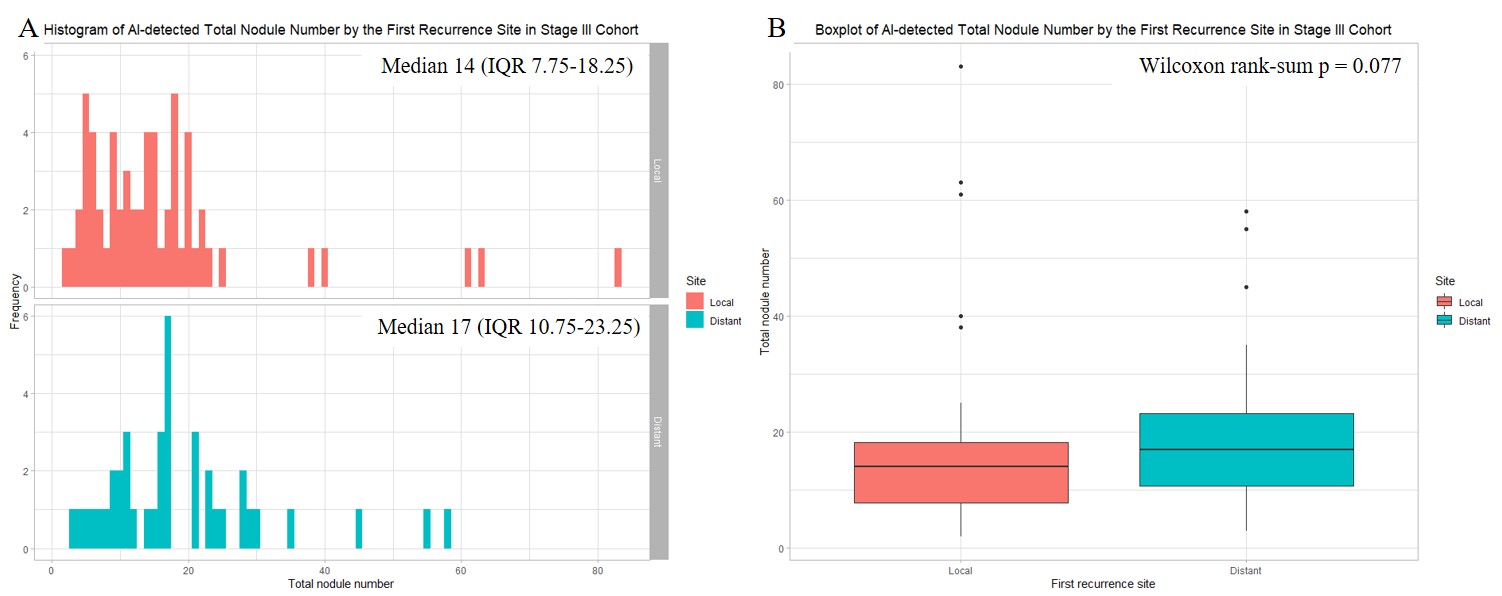

**Supplementary Figure 4. AI-detected TNN stratified by recurrence pattern in stage III cohort.** (A) histograms showing the frequency distribution, (B) boxplots showing the central tendency. TNN, total nodule number.

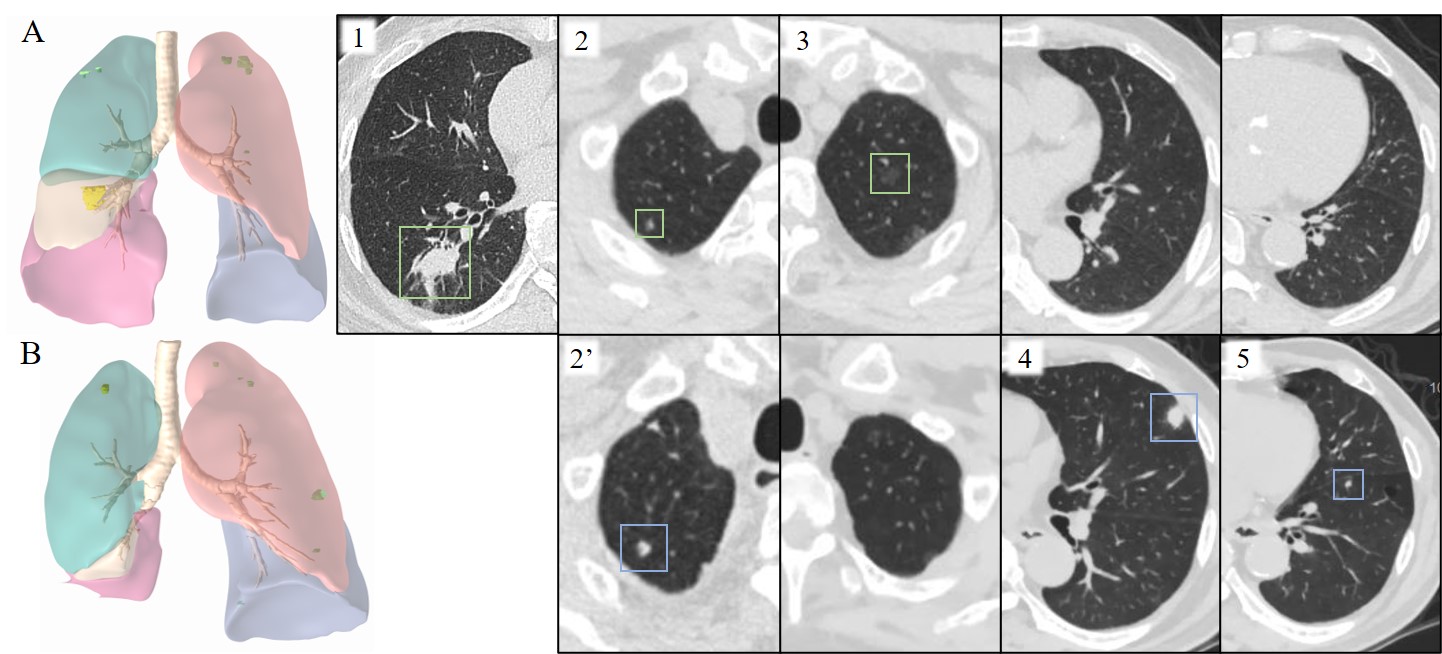

**Supplementary Figure 5. A case report: 80-year-old male underwent right lower lobe lobectomy and diagnosed with pathologic stage IIIA lung adenocarcinoma.** (A) Preoperative chest CT showed the primary tumor at right lower lobe (#1); a small solid nodule with benign-looking radiological features at right upper lobe (#2), which also exhibited no FDG uptake on PET-CT; and a pure GGN at left upper lobe (#3). (B) Postoperative follow-up chest CT showed the growth of right upper lobe nodule (#2 to #2’), which indicated its malignancy; and the occurrence of new solid nodules at left upper lobe (#4) and left lower lobe (#5), which suggested intrapulmonary metastases. FDG, fluorodeoxyglucose; PET, positron emission tomography; GGN, ground-glass nodule.
